# A Scoping Review of Factors used to Explain Disparities in COVID-19 Vaccination Intentions and Uptake among People of Color—United States, December 1, 2020-April 30, 2021

**DOI:** 10.1101/2023.01.12.23284499

**Authors:** Rebecca F. Wilson, Krishna Kiran Kota, Kameron J. Sheats, Carolina Luna-Pinto, Chantelle Owens, Dominique D. Harrison, Sima Razi

## Abstract

**Background:** Vaccine access, coupled with the belief that vaccines are important, beneficial, and safe, plays a pivotal role in achieving high levels of vaccination to reduce the spread and severity of COVID-19 in the United States (U.S.) and globally. Many factors can influence vaccine intentions and uptake.

**Methods:** We conducted a scoping review of factors (e.g., access-related factors, racism) known to influence vaccine intentions and uptake, using publications from various databases and websites published December 1, 2020-April 30, 2021. Descriptive statistics were used to present results.

**Results:** Overall, 1094 publications were identified through the database search, of which 133 were included in this review. Among the publications included, over 60% included mistrust in vaccines and vaccine-safety concerns, 43% included racism/discrimination, 35% included lack of vaccine access (35%), and 8% had no contextual factors when reporting on vaccine intentions and disparities in vaccine uptake.

**Conclusions:** Findings revealed during a critical period when there was a well-defined goal for adult COVID-19 vaccination in the U.S., some publications included several contextual factors while others provided limited or no contextual factors when reporting on disparities in vaccine intentions and uptake. Failing to contextualize inequities and other factors that influence vaccine intentions and uptake might be perceived as placing responsibility for vaccination status on the individual, consequently, leaving social and structural inequities that impact vaccination rates and vaccine confidence, among people of color, intact.

## INTRODUCTION

Vaccine access, coupled with the belief that vaccines are important, beneficial, and safe, plays a pivotal role in achieving high levels of vaccination to reduce the spread and severity of COVID-19 in the United States (U.S.) and globally.^1,2^ The delay in acceptance or refusal of vaccination despite availability of vaccination services, called vaccine hesitancy, is understood as a continuum between those who totally accept and those who refuse all vaccines.^3^After the December 2020 rollout of COVID-19 vaccines in the U.S., overcoming vaccine hesitancy, seen as a critical pathway to mitigating the impact of COVID-19, has been examined extensively within the context of vaccination intentions and vaccine uptake among people of color.^4-6^ Many factors can influence vaccine acceptance and uptake, including inequitable access to vaccines, experiences with racism and discrimination, mistrust in systems (e.g., medical establishments, government) responsible for equitable outcomes, vaccine hesitancy, language barriers, etc.^7-12^ The purpose of this scoping review is to examine the frequency with which publications included contextual factors when using the term vaccine hesitancy or its synonyms to report disparities in COVID-19 vaccine intentions and uptake among people of color during the early stages of the rollout of COVID-19 vaccines.

## METHODS

This study uses a scoping review, which is a rigorous methodological approach used for purposes such as summarizing and disseminating research findings, identifying research gaps, and making recommendations for future research.^13,14^

### Eligibility criteria

The inclusion criteria for the scoping review were: 1) news articles and stories, journal articles, and grey literature (e.g., working papers) that: 2) were published in English, 3) used vaccine hesitancy-related terminology (see Supplementary Table 1) to report racial and ethnic disparities in vaccine intentions and uptake in the U.S. (including U.S. territories), and 4) were published December 1, 2020 through April 30, 2021. The starting date coincided with the December 2020 U.S. initial rollout of COVID-19 vaccines, and the cutoff date aligned with President Biden’s deadline of April 30, 2021 for states to expand eligibility of COVID-19 vaccines to all adults.^15^ The review excluded blogs, podcasts, websites, audio and video clips, and transcripts to allow for a more focused scoping review, as these sources tend to report on a broader range of topics, beyond disparities in vaccine uptake and intentions.

**Table 1:**
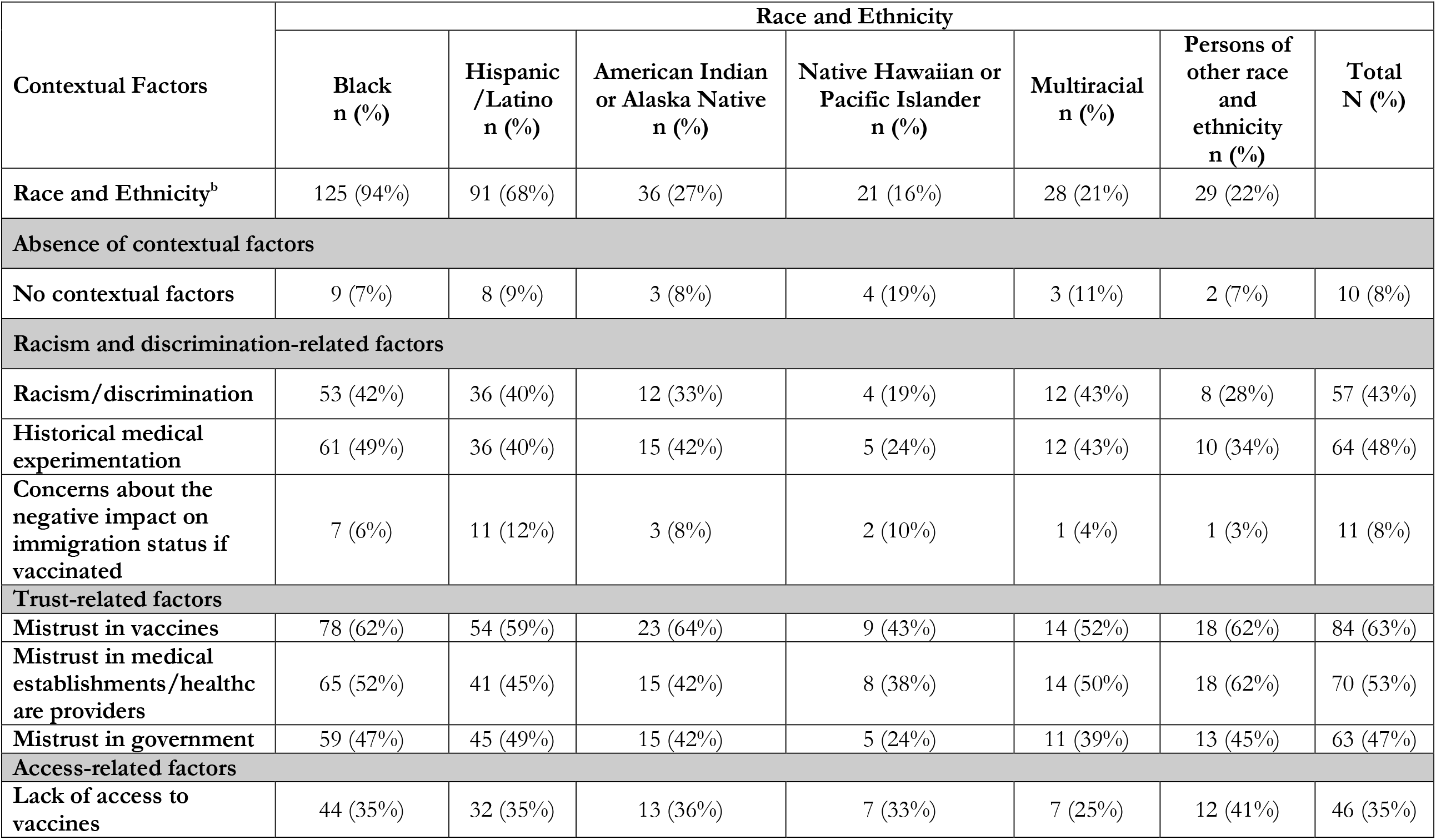
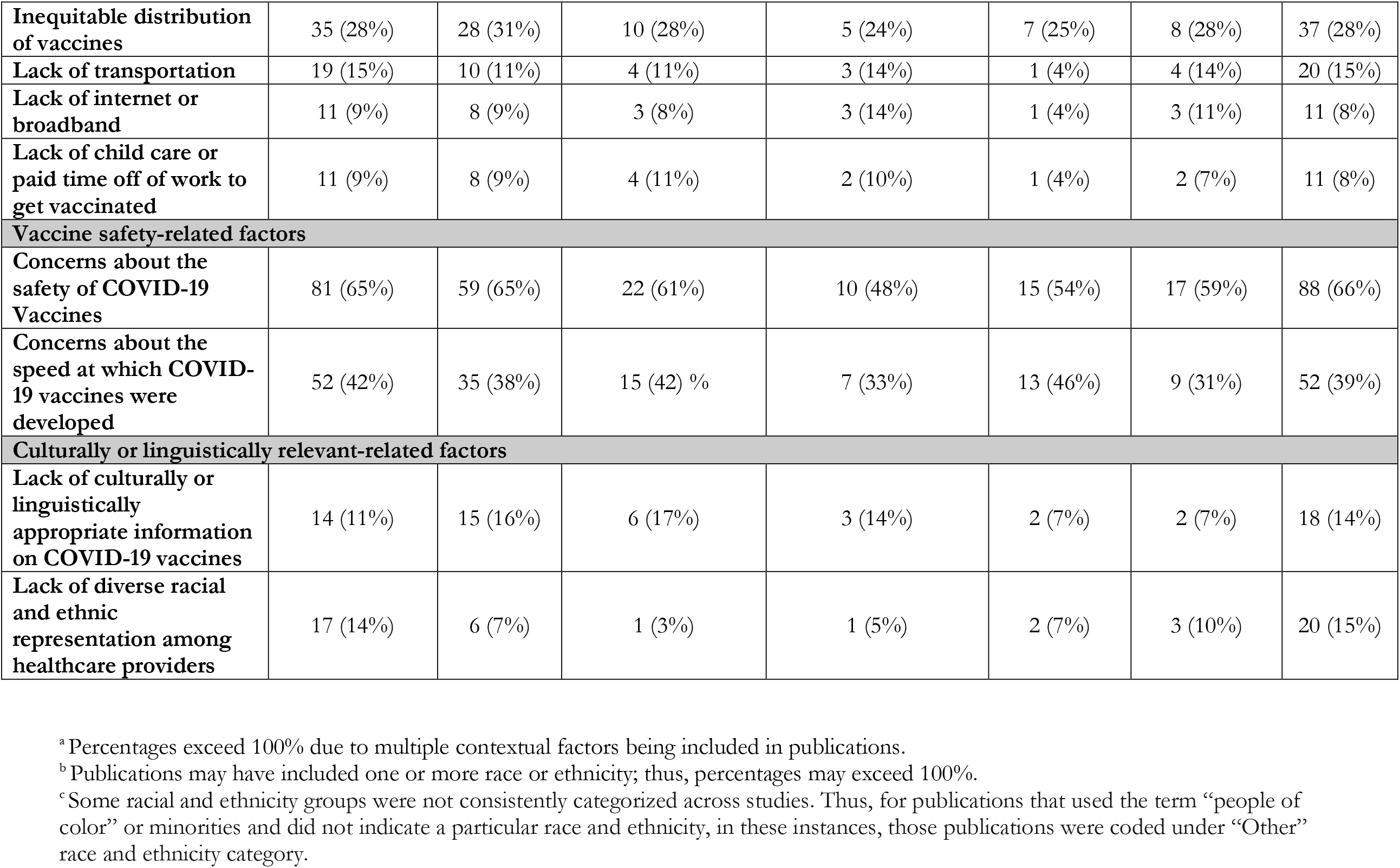
Number and Percentages^a^ of Contextual factors by race and ethnicity^b^ of People of Color^c^ extracted from Publications in Mediums of Communication (N=133)

| Contextual Factors | Race and Ethnicity |  |  |  |  |  |  |
| --- | --- | --- | --- | --- | --- | --- | --- |
|  | Black<br>n (%) | Hispanic<br>/Latino<br>n (%) | American Indian<br>or Alaska Native<br>n (%) | Native Hawaiian or<br>Pacific Islander<br>n (%) | Multiracial<br>n (%) | Persons of<br>other race<br>and<br>ethnicity<br>n (%) | Total<br>N (%) |
| Race and Ethnicity<sup>b</sup> | 125 (94%) | 91 (68%) | 36 (27%) | 21 (16%) | 28 (21%) | 29 (22%) |  |
| **Absence of contextual factors** |  |  |  |  |  |  |  |
| No contextual factors | 9 (7%) | 8 (9%) | 3 (8%) | 4 (19%) | 3 (11%) | 2 (7%) | 10 (8%) |
| **Racism and discrimination-related factors** |  |  |  |  |  |  |  |
| Racism/discrimination | 53 (42%) | 36 (40%) | 12 (33%) | 4 (19%) | 12 (43%) | 8 (28%) | 57 (43%) |
| Historical medical experimentation | 61 (49%) | 36 (40%) | 15 (42%) | 5 (24%) | 12 (43%) | 10 (34%) | 64 (48%) |
| Concerns about the negative impact on immigration status if vaccinated | 7 (6%) | 11 (12%) | 3 (8%) | 2 (10%) | 1 (4%) | 1 (3%) | 11 (8%) |
| **Trust-related factors** |  |  |  |  |  |  |  |
| Mistrust in vaccines | 78 (62%) | 54 (59%) | 23 (64%) | 9 (43%) | 14 (52%) | 18 (62%) | 84 (63%) |
| Mistrust in medical establishments/healthcare providers | 65 (52%) | 41 (45%) | 15 (42%) | 8 (38%) | 14 (50%) | 18 (62%) | 70 (53%) |
| Mistrust in government | 59 (47%) | 45 (49%) | 15 (42%) | 5 (24%) | 11 (39%) | 13 (45%) | 63 (47%) |
| **Access-related factors** |  |  |  |  |  |  |  |
| Lack of access to vaccines | 44 (35%) | 32 (35%) | 13 (36%) | 7 (33%) | 7 (25%) | 12 (41%) | 46 (35%) |
| **Inequitable distribution of vaccines** | 35 (28%) | 28 (31%) | 10 (28%) | 5 (24%) | 7 (25%) | 8 (28%) | 37 (28%) |
| **Lack of transportation** | 19 (15%) | 10 (11%) | 4 (11%) | 3 (14%) | 1 (4%) | 4 (14%) | 20 (15%) |
| **Lack of internet or broadband** | 11 (9%) | 8 (9%) | 3 (8%) | 3 (14%) | 1 (4%) | 3 (11%) | 11 (8%) |
| **Lack of child care or paid time off of work to get vaccinated** | 11 (9%) | 8 (9%) | 4 (11%) | 2 (10%) | 1 (4%) | 2 (7%) | 11 (8%) |
| **Vaccine safety-related factors** |  |  |  |  |  |  |  |
| **Concerns about the safety of COVID-19 Vaccines** | 81 (65%) | 59 (65%) | 22 (61%) | 10 (48%) | 15 (54%) | 17 (59%) | 88 (66%) |
| **Concerns about the speed at which COVID-19 vaccines were developed** | 52 (42%) | 35 (38%) | 15 (42) % | 7 (33%) | 13 (46%) | 9 (31%) | 52 (39%) |
| **Culturally or linguistically relevant-related factors** |  |  |  |  |  |  |  |
| **Lack of culturally or linguistically appropriate information on COVID-19 vaccines** | 14 (11%) | 15 (16%) | 6 (17%) | 3 (14%) | 2 (7%) | 2 (7%) | 18 (14%) |
| **Lack of diverse racial and ethnic representation among healthcare providers** | 17 (14%) | 6 (7%) | 1 (3%) | 1 (5%) | 2 (7%) | 3 (10%) | 20 (15%) |

<sup>a</sup> Percentages exceed 100% due to multiple contextual factors being included in publications.
<sup>b</sup> Publications may have included one or more race or ethnicity; thus, percentages may exceed 100%.
<sup>c</sup> Some racial and ethnicity groups were not consistently categorized across studies. Thus, for publications that used the term “people of color” or minorities and did not indicate a particular race and ethnicity, in these instances, those publications were coded under “Other” race and ethnicity category.

### Search

With the assistance of a reference librarian, the study team searched the following databases on May 20, 2021: US Newsstream, Medline, Embase, PsychInfo, Global Health, CINAHL, Academic Search Complete, Coronavirus Research Database, SCOPUS, and World Health Organization COVID-19 Literature databases and websites. The search terms focused on COVID-19 vaccine hesitancy and people of color. The full search strategy and terms are available in Appendix 1. The search yielded newspaper articles, commentaries and editorials, academic peer-reviewed journal articles, and grey literature, including working papers and preprints.

### Data extraction and synthesis

Each publication was assigned two reviewers (CL-P, CO, KK, and RFW) who independently examined the publications to determine eligibility for inclusion in the study. Discrepancies were reconciled by at least two abstractors until full consensus was reached. Reviewers coded the publications for contextual factors (i.e., racism and discrimination-related factors, trust-related factors, access-related factors, vaccine safety-related factors, culturally and linguistically relevant-related factors, and absence of contextual factors; Table 1). Reviewers also coded racial and ethnicity groups mentioned within the context of vaccine hesitancy. Analysis used descriptive statistics to summarize contextual factors and race and ethnicity included in publications.

### Data items

The contextual factors (Table 1), were grouped into the following six broad categories: *racism and discrimination-related factors* (racism/discrimination, historical medical experimentation, concerns about change in immigration status); *trust-related factors* (mistrust in vaccines, mistrust in medical establishment(s)/healthcare providers, mistrust in government); *access-related factors* (lack of access to vaccines, inequitable distribution of vaccine, lack of transportation, lack of internet or broadband, lack of child care or paid time off of work); *vaccine safety-related factors* (concerns about vaccine safety, concerns about speed at which COVID-19 vaccines were developed); *culturally relevant-related factors* (lack of culturally or linguistically appropriate information on COVID-19 vaccines, lack of racial and ethnic representation among those promoting or administering COVID-19 vaccines); and *no contextual factors*. The lack of contextual factors (*no contextual factors*) is examined to highlight the absence of information that could provide important context about vaccine intentions and uptake among people of color.

## RESULTS

### Racial and ethnic groups

From December 1, 2020 through April 30, 2021, a total of 133 publications were reviewed and included in this report. Two-thirds of the publications were journal articles (68%), followed by news articles (28%), and trade journal articles (4%) (Figure 1). The majority of publications mentioned Black persons (94%), followed by Hispanic or Latino (68%), American Indian or Alaska Native (27%), Native Hawaiian or Pacific Islander (16%), multiracial (21%), and other race and ethnicity (22%) persons.

**Figure 1.**
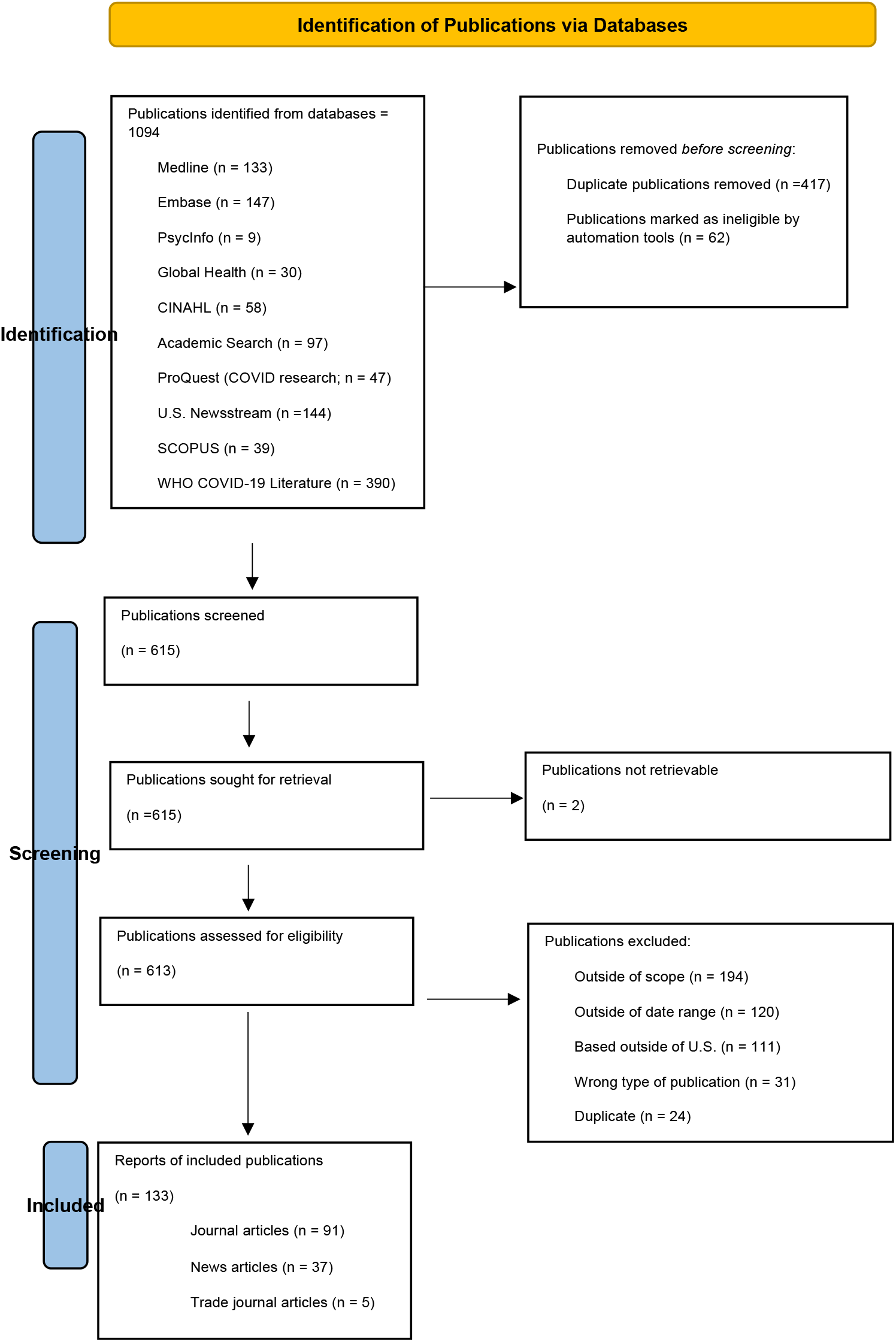
Flowchart of Study Selection. **Abbreviations:** U.S. (United States); WHO (World Health Organization) Overall, 1094 publications were identified through the database search (Figure 1), of which 479 were removed before screening. Among the 615 publications screened, 2 were not retrievable, leaving 613 to be assessed for eligibility. Of the 613 publications screened for eligibility, 480 were excluded, and 133 were included in this scoping review. Reasons for exclusion were as follows: outside of scope of the study (n=194), outside date range (n=120), based outside of the U.S. (n=111), publication type not eligible (n=31), and duplicate record (n=24).

### Select contextual factors by race and ethnicity

#### Black persons

Of publications (94%; n=125) that reported on disparities in vaccine intentions and uptake among Black persons in the U.S., 7% provided zero contextual factors. Less than half (42%) included racism/discrimination, and 49% included historical medical experimentation. Mistrust in vaccines was mentioned in 62% of publications. The lack of access to vaccines and concerns about vaccine safety were mentioned in 35% and 65% of publications, respectively. The lack of racial and ethnic representation among healthcare providers was included in 14% of publications.

#### Hispanic or Latino persons

Hispanic or Latino persons were included in 68% (n=91) of the 133 publications, of which 9% (n=8) did not provide any contextual factors. Racism/discrimination and historical medical experimentation were each mentioned in 40% of publications, whereas concerns about the potential negative impact on immigration status if accessing vaccination services was mentioned in 12% of publications. Mistrust in vaccines was included in 59% of publications. Lack of access to vaccines and concerns about the safety of vaccines were mentioned in 35% and 65% of publications, respectively. Lack of culturally or linguistically appropriate information on COVID-19 vaccines was included in 16% of publications.

#### American Indian or Alaska Native persons

Slightly more than one-quarter (27%; n=36) of the publications mentioned American Indian or Alaska Native persons. Of them, 8% (n=3) provided zero contextual factors, 33% included racism/discrimination, and 42% included historical medical experimentation as contextual factors. Mistrust in vaccines and lack of access to vaccines were included in 64% and 36% of publications, respectively. Over half (61%) of publications reported concerns about vaccine safety, and 17% included lack of culturally or linguistically appropriate information.

#### Native Hawaiian or Pacific Islander persons

Twenty-one (16%) publications included Native Hawaiian or Pacific Islander persons. Of them, 19% provided zero contextual factors. Almost one-fifth (19%) included racism/discrimination. Mistrust in vaccines and mistrust in healthcare providers were included in 43% and 38% of publications, respectively. Lack of access to vaccines was included in 33% of publications. Concerns about vaccine safety was included in 48% of publications, and 14% included lack of culturally or linguistically appropriate information on COVID-19 vaccines.

### Limitations

While this scoping review contributes to the literature and provides a rapid review of the extent to which publications contextualized factors that impacted earlier disparities in COVID-19 vaccine intentions and uptake among people of color, several limitations must be acknowledged. First, this scoping review did not evaluate the quality or depth in which media (e.g., scientific research, news media, newspapers) contextualized factors, but coded for the presence and absence of contextual factors. There might have been instances where contextual factors were mentioned and not expanded upon, possibly leading to a misrepresentation of how well-represented contextual factors were in publications. Second, examining pre-specified contextual factors may have led to the exclusion of other contextual factors. Third, some publications may have conflated some contextual factors, making it challenging for us to disentangle which contextual factor to code; this may have led to under and over-counting some contextual factors. Fourth, scoping reviews are at risk of bias; thus, there may have been undetected instances where bias was introduced. We sought to remove potential bias by having two abstractors independently review each publication and reconcile all discrepancies until 100% consensus was reached. Fifth, we relied on the assistance of a reference librarian to provide the initial list of publications utilizing specified search terms; selection bias may have occurred if the search strategy did not identify all available publications on vaccine hesitancy, resulting in the exclusion of some publications. Sixth, the focus of trade journals, news articles, and academic peer-reviewed articles varies; thus, the number of contextual factors present in these different types of publications may vary. Seventh, this scoping review only included publications that were published December 1, 2020-April 30, 2021, which was early in the vaccine roll out. Since then, using a health equity approach, substantial strides have been made to reducing racial and ethnic differences in vaccine coverage.^16-18^

## Conclusions

Despite these limitations, this scoping review highlights the fact that during the initial stages of vaccine rollout, in this review, some publications failed to contextualize how certain factors (e.g., racism, structural and social inequities) might influence vaccine-seeking behaviors among people of color during a critical early period in vaccine rollout. Further, in situations where vaccine intentions and uptake are not attributed to hesitancy, focusing on “refusal” and not inequities, may propagate stigmatizing views and harmful narratives about people of color, undermining vaccination efforts.

## Data Availability

All data produced in the present study are available upon reasonable request to the authors.

## Appendix 1 Search Query

A Scoping Review of Factors used to Explain Disparities in COVID-19 Vaccination Intentions and Uptake among People of Color—United States, December 1, 2020-April 30, 2021

This search strategy consisted of the following terms or their derivative: vaccine hesitancy, vaccine intentions, vaccine confidence, vaccine attitude, vaccine belief, vaccine refusal, vaccine skepticism, vaccine outreach, vaccine knowledge, vaccine practice, vaccine attitude, vaccine belief, vaccine resistance, vaccine stigma, and vaccine choice/decision in combination with the following terms or their derivative: race, racial, ethnic, disparity, equitable, communities of color, minorities, African American, Black people, Black Americans, Black community, brown people, brown Americans, brown communities, Hispanic, Latino, Latina, Latinx, Pacific Islander, Native American, American Indian, Alaska Native, Tribal communities, or tribal population.

| Database | Search Strategy | Key Themes |
| --- | --- | --- |
| **Medline (OVID) 1946-** | (Vaccin* ADJ5 (intent* OR intend* OR confiden* OR hesitan* OR outreach OR knowledge OR practice* OR attitude* OR belief* OR accept* OR choice OR decision* OR refusal OR refuse OR skeptic* OR resist* OR stigma* OR rate*))<br>AND<br>Race OR racial OR ethnic* OR disparit* OR equit* OR (communit* ADJ2 color) OR minorit* OR African American* OR (Black ADJ2 (people OR american* OR communit*) OR (brown ADJ2 (people OR american* OR communit*) OR Hispanic* OR Latino* OR Latina* OR Latinx OR Native American* OR American Indian* OR AI?NA OR Alaska Native* OR Tribal communit* OR tribal population*<br>Limit to COVID-19<br>December 2020 - April 2021 | Vaccine hesitancy, people of color, COVID-19 vaccine |
| **Embase (OVID) 1974-** | (Vaccin* ADJ5 (intent* OR intend* OR confiden* OR hesitan* OR outreach OR knowledge OR practice* OR attitude* OR belief* OR accept* OR choice OR decision* OR refusal OR refuse OR skeptic* OR resist* OR stigma* OR rate*))<br>AND<br>Race OR racial OR ethnic* OR disparit* OR equit* OR (communit* ADJ2 color) OR minorit* OR African American* OR (Black ADJ2 (people OR american* OR communit*) OR (brown ADJ2 (people OR american* OR communit*) OR Hispanic* OR Latino* OR Latina* OR Latinx OR Native American* OR American Indian* OR AI?NA OR Alaska Native* OR Tribal communit* OR tribal population*<br>Limit to COVID-19 | Vaccine hesitancy, people of color, COVID-19 vaccine |
|  | December 2020 - April 2021 |  |
| **PsycInfo**<br>**(OVID)**<br>**1987-** | (novel coronavir* OR novel corona virus* OR 2019 coronavirus OR betacoronavir* OR covid19 OR covid 19 OR nCoV OR novel CoV OR CoV 2 OR CoV2 OR sarscov2 OR sars-cov OR sarscov OR 2019nCoV OR 2019-nCoV OR coronavirus infections)<br>AND<br>(Vaccin* ADJ5 (intent* OR intend* OR confiden* OR hesitan* OR outreach OR knowledge OR practice* OR attitude* OR belief* OR accept* OR choice OR decision* OR refusal OR refuse OR skeptic* OR resist* OR stigma* OR rate*))<br>AND<br>Race OR racial OR ethnic* OR disparit* OR equit* OR (communit* ADJ2 color) OR minorit* OR African American* OR (Black ADJ2 (people OR american* OR communit*)) OR (brown ADJ2 (people OR american* OR communit*)) OR Hispanic* OR Latino* OR Latina* OR Latinx OR Native American* OR American Indian* OR AI?NA OR Alaska Native* OR Tribal communit* OR tribal population*<br>December 2020 - April 2021 | Vaccine<br>hesitancy,<br>people of<br>color,<br>COVID-19<br>vaccine |
| **Global**<br>**Health**<br>**OVID** | (novel coronavir* OR novel corona virus* OR 2019 coronavirus OR betacoronavir* OR covid19 OR covid 19 OR nCoV OR novel CoV OR CoV 2 OR CoV2 OR sarscov2 OR sars-cov OR sarscov OR 2019nCoV OR 2019-nCoV OR coronavirus infections)<br>AND<br>(Vaccin* ADJ5 (intent* OR intend* OR confiden* OR hesitan* OR outreach OR knowledge OR practice* OR attitude* OR belief* OR accept* OR choice OR decision* OR refusal OR refuse OR skeptic* OR resist* OR stigma* OR rate*))<br>AND<br>Race OR racial OR ethnic* OR disparit* OR equit* OR (communit* ADJ2 color) OR minorit* OR African American* OR (Black ADJ2 (people OR american* OR communit*)) OR (brown ADJ2 (people OR american* OR communit*)) OR Hispanic* OR Latino* OR Latina* OR Latinx OR Native American* OR American Indian* OR AI?NA OR Alaska Native* OR Tribal communit* OR tribal population*<br>December 2020 - April 2021 | Vaccine<br>hesitancy,<br>people of<br>color,<br>COVID-19<br>vaccine |
| **CINAHL**<br>**(EbscoHost)** | (“novel coronavir*” OR “novel corona virus*” OR “2019 coronavirus” OR betacoronavir* OR covid19 OR “covid 19” OR nCoV OR “novel CoV” OR “CoV 2” OR CoV2 OR sarscov2 OR sars-cov OR sarscov OR 2019nCoV OR 2019-nCoV OR “coronavirus infections”)<br>AND | Vaccine<br>hesitancy,<br>people of<br>color, |
|  | (Vaccin* N5 (intent* OR intend* OR confiden* OR hesitan* OR outreach OR knowledge OR practice* OR attitude* OR belief* OR accept* OR choice OR decision* OR refusal OR refuse OR skeptic* OR resist* OR stigma* OR rate*))<br>AND<br>Race OR racial OR ethnic* OR disparit* OR equit* OR (communit* N2 color) OR minorit* OR "African American*" OR (Black N2 (people OR american* OR communit*)) OR (brown N2 (people OR american* OR communit*)) OR Hispanic* OR Latino* OR Latina* OR Latinx OR "Native American*" OR "American Indian*" OR AI?NA OR "Alaska Native*" OR "Tribal communit*" OR "tribal population*"<br>December 2020 - April 2021 | COVID-19 vaccine |
| **Academic Search Complete** | ("novel coronavir*" OR "novel corona virus*" OR "2019 coronavirus" OR betacoronavir* OR covid19 OR "covid 19" OR nCoV OR "novel CoV" OR "CoV 2" OR CoV2 OR sarscov2 OR sars-cov OR sarscov OR 2019nCoV OR 2019-nCoV OR "coronavirus infections")<br>AND<br>(Vaccin* N5 (intent* OR intend* OR confiden* OR hesitan* OR outreach OR knowledge OR practice* OR attitude* OR belief* OR accept* OR choice OR decision* OR refusal OR refuse OR skeptic* OR resist* OR stigma* OR rate*))<br>AND<br>Race OR racial OR ethnic* OR disparit* OR equit* OR (communit* N2 color) OR minorit* OR "African American*" OR (Black N2 (people OR american* OR communit*)) OR (brown N2 (people OR american* OR communit*)) OR Hispanic* OR Latino* OR Latina* OR Latinx OR "Native American*" OR "American Indian*" OR AI?NA OR "Alaska Native*" OR "Tribal communit*" OR "tribal population*"<br>December 2020 - April 2021 | Vaccine hesitancy, people of color, COVID-19 vaccine |
| **Coronavirus Research Database (ProQuest)** | TI,AB("novel coronavir*" OR "novel corona virus*" OR "2019 coronavirus" OR betacoronavir* OR covid19 OR "covid 19" OR nCoV OR "novel CoV" OR "CoV 2" OR CoV2 OR sarscov2 OR sars-cov OR sarscov OR 2019nCoV OR 2019-nCoV OR "coronavirus infections")<br>AND<br>TI,AB(Vaccin* NEAR/5 (intent* OR intend* OR confiden* OR hesitan* OR outreach OR knowledge OR practice* OR attitude* OR belief* OR accept* OR choice OR decision* OR refusal OR refuse OR skeptic* OR resist* OR stigma* OR rate*))<br>AND | Vaccine hesitancy, people of color, COVID-19 vaccine |
|  | TI,AB(Race OR racial OR ethnic* OR disparit* OR equit* OR (communit* NEAR/2 color) OR minorit* OR "African American*" OR (Black NEAR/2 (people OR american* OR communit*)) OR (brown NEAR/2 (people OR american* OR communit*)) OR Hispanic* OR Latino* OR Latina* OR Latinx OR "Native American*" OR "American Indian*" OR AI?NA OR "Alaska Native*" OR "Tribal communit*" OR "tribal population*")<br>December 2020 - April 2021 |  |
| **US Newsstream**<br>**ProQuest Central** | TI,AB("novel coronavir*" OR "novel corona virus*" OR "2019 coronavirus" OR betacoronavir* OR covid19 OR "covid 19" OR nCoV OR "novel CoV" OR "CoV 2" OR CoV2 OR sarscov2 OR sars-cov OR sarscov OR 2019nCoV OR 2019-nCoV OR "coronavirus infections")<br>AND<br>TI,AB(Vaccin* NEAR/5 (intent* OR intend* OR confiden* OR hesitan* OR outreach OR knowledge OR practice* OR attitude* OR belief* OR accept* OR choice OR decision* OR refusal OR refuse OR skeptic* OR resist* OR stigma* OR rate*))<br>AND<br>TI,AB(Race OR racial OR ethnic* OR disparit* OR equit* OR (communit* NEAR/2 color) OR minorit* OR "African American*" OR (Black NEAR/2 (people OR american* OR communit*)) OR (brown NEAR/2 (people OR american* OR communit*)) OR Hispanic* OR Latino* OR Latina* OR Latinx OR "Native American*" OR "American Indian*" OR AI?NA OR "Alaska Native*" OR "Tribal communit*" OR "tribal population*")<br>December 2020 - April 2021 | Vaccine hesitancy, people of color, COVID-19 vaccine |
| **Scopus** | TITLE-ABS-KEY("novel coronavir*" OR "novel corona virus*" OR "2019 coronavirus" OR betacoronavir* OR covid19 OR "covid 19" OR nCoV OR "CoV 2" OR cov2 OR sarscov2 OR sars-cov OR sarscov OR 2019nCoV OR 2019-nCoV OR "novel CoV" OR "coronavirus infections") AND TITLE-ABS-KEY(Vaccin* W/5 (intent* OR intend* OR confiden* OR hesitan* OR outreach OR knowledge OR practice* OR attitude* OR belief* OR accept* OR choice OR decision* OR refusal OR refuse OR skeptic* OR resist* OR stigma* OR rate*)) AND TITLE-ABS-KEY(Race OR racial OR ethnic* OR disparit* OR equit* OR (communit* W/2 color) OR minorit* OR "African American*" OR (Black W/2 (people OR american* OR communit*)) OR (brown W/2 (people OR american* OR communit*)) OR Hispanic* OR Latino* OR Latina* OR Latinx OR "Native American*" OR "American Indian*" OR AI?NA OR "Alaska Native*" OR "Tribal communit*" OR "tribal population*") AND NOT INDEX(medline)<br>December 2020 - April 2021 | Vaccine hesitancy, people of color, COVID-19 vaccine |
| **WHO**<br>**COVID-19**<br>**Literature** | Vaccin* AND (intention OR intend* OR confiden* OR hesitan* OR outreach OR attitude OR belief* OR accept* OR choice OR decision* OR refusal OR refuse OR skeptic* OR resist* OR skeptic* OR resist* OR stigma* OR rate*)<br>AND<br>Race OR racial OR ethnic* OR disparit* OR equit* OR "communities of color" OR "community of color" OR minorit* OR "African American" OR "African Americans" OR "Black people" OR "Black American" OR "black americans" OR "black communities" OR "black community" OR "Brown people" OR "Brown American" OR "brown americans" OR "brown communities" OR "brown community" OR Hispanic OR Latino OR Latina OR Hispanics OR Latinos OR Latinas OR Latinx OR "Native American" OR "American Indian" OR "Native Americans" OR "American Indians" OR "Alaska Native" OR "Alaska Natives" OR Tribal<br>December 2020 - April 2021 | Vaccine<br>hesitancy,<br>people of<br>color,<br>COVID-19<br>vaccine |

